# Detection of dynamic lung hyperinflation using cardiopulmonary exercise testing and respiratory function in patients with stable cardiac disease: a multicenter cross-sectional study

**DOI:** 10.1101/2023.04.27.23289236

**Authors:** Kazuyuki Kominami, Kazuki Noda, Nanaho Minagawa, Kazuya Yonezawa, Masanori Ueda, Yasuyuki Kobayashi, Makoto Murata, Masatoshi Akino

## Abstract

**Background:** Many patients with heart disease potentially have comorbid COPD, however there are not enough opportunities for screening and the qualitative differentiation of shortness of breath (SOB) has not been well established. We investigated the detection rate of SOB based on a visual and qualitative dynamic lung hyperinflation (DLH) detection index during cardiopulmonary exercise testing (CPET) and whether there was a difference in respiratory function between the two groups.

**Methods:** We recruited 534 patients with heart disease and to scrutinize physical functions (369 males, 67.0±12.9 years) who underwent CPET and spirometry simultaneously. The difference between inspiratory and expiratory tidal volume was calculated (TV E-I) from the breath-by-breath data. A TV E-I decrease after the start of exercise comprised the convex group, and a TV E-I that remained unchanged or increased comprised the non-convex group.

**Results:** 129 patients (24.2%) were in the convex; there was no difference in clinical characteristics between groups. The Borg scale scores at the end of the CPET showed no difference. VE/VCO2 slope, its Y-intercept and minimum VE/VCO2 showed no significant difference. In the convex group, FEV1.0% was significantly lower (69.4±13.1 vs. 75.0±9.0%), and FEV1.0% and Y-intercept and the difference between minimum VE/VCO2 and VE/VCO2 slope were significantly correlated (r=−0.343 and r=−0.478).

**Conclusions:** The convex group showed decreased respiratory function, suggesting potential airway obstruction during exercise. A combined assessment of the TV E-I and Y-intercept of the VE/VCO2 slope or the difference between the minimum VE/VCO2 and VE/VCO2 slopes could potentially detect COPD or airway obstruction.

## Introduction

Patients with cardiovascular diseases usually suffer from respiratory diseases such as chronic obstructive pulmonary disease (COPD) [1–3], and develop fatigue and shortness of breath because of a variety of factors that limit exercise and activity, including lifestyle factors such as inactivity [4], increased left ventricular filling pressures [5], ventilatory-perfusion mismatch [6], and impaired oxygen delivery capacity because of cardiac dysfunction [7]. Current smoking or ex-smoking are common coronary risk factors and exacerbating factors of chronic heart failure [8–12]. In addition, poor lung function is an independent risk factor for cardiovascular disease and atrial fibrillation (AF) [13–17], and subclinical respiratory impairment is associated with the development of hypertension, which is a major risk factor for cardiovascular disease and mortality [18]. However, comorbid COPD without a history of smoking [19], potentially comorbid COPD, or respiratory impairment may not be adequately assessed, and this has not led to early detection of COPD or respiratory impairment in patients with heart disease [20].

Airflow obstruction in COPD is caused by decreased pulmonary elastic contractile pressure because of peripheral airway involvement and emphysematous lesions, resulting in collapsed airways and air trapped in the lungs during forced expiration (referred to as “air trapping”). Collapsed peripheral airways also occur during resting breathing as the disease progresses, contributing to lung hyperinflation [21]. In addition, air trapping, caused by collapsed peripheral airways, is strengthened with exertion or exercise, causing further lung hyperinflation. This is called dynamic lung hyperinflation (DLH) and is an important factor in patients with COPD, contributing to increased respiratory workload, shortness of breath on exertion, and reduced exercise tolerance [22, 23]. Although measurement of inspiratory capacity (IC) during exercise testing or the hyperventilation method has been proposed to assess DLH [24–29], opportunities for screening are insufficient. In contrast, there have been several reports combining exercise testing and inspiratory reserve capacity; however, there are limited reports on cardiopulmonary exercise testing (CPET), which is the most standard method of assessing exercise tolerance [30–35]. Furthermore, CPET, the gold standard for assessing exercise tolerance, provides information on the index of exercise tolerance. In CPET, the Y-intercept of the linear carbon dioxide production (CO_2_) and minute ventilation (VE) relationship (the VE/VCO_2_ slope) is related to the severity of COPD and the forced expiratory volume (FEV) in 1 s (FEV1.0) as a percentage of forced vital capacity (FEV 1.0%) [36]. In previous studies, we have attempted to use CPET measurements to visually and qualitatively detect the DLH [37]. In this method, the difference between the expiratory and inspiratory tidal volumes is measured during incremental exercise, and expiration is assumed to be reduced relative to inspiration when dynamic lung hyperinflation occurs.

In this study, we hypothesized that the presence or absence of the DLH index would be correlated with the CPET indices and respiratory function. By clarifying the relationship between the presence or absence of the indicators of DLH, CPET, and respiratory function, we believe a detailed evaluation of respiratory function using CPET is possible. In addition, identifying respiratory impairment by CPET in asymptomatic or very mild stages of COPD will permit early preventive intervention and help prevent disease exacerbation.

## Methods

We included 534 patients with stable heart disease and to scrutinize physical functions (age:67.0 ± 12.9 years [95%CI: 65.9–68.1], height:161.9 ± 9.2 cm [95%CI: 161.2–162.7], body weight:62.5 ± 14.6 kg [95%CI: 61.3–63.7], BMI:23.7 ± 4.4 kg/m^2^ [95%CI: 23.3–24.1]) who underwent CPET and spirometry testing approximately the same time.

The CPET was performed using a stationary bicycle (StrengthErgo 8; Mitsubishi Electric Engineering, Tokyo, COMBI 75XL3; Konami Sports Co., Ltd., Tokyo) on a breath-by-breath basis with a gas analyzer (AE-300S or AE-310S; Minato Ikagaku Co., Tokyo). The maximal symptomatic exercise was performed using the ramp protocol. The exercise protocol consisted of 2–3 minutes of rest and 2–3 minutes of warm-up (W.U). The ramp protocol was adjusted to 10–20 W/min, assuming the individual exercise tolerance level. The rating of perceived exertion (RPE) at the end of the exercise was assessed using the Borg scale.

Furthermore, a breath-by-breath gas analyzer (AE-300S or AE-310S; Minato Ikagaku Co., Tokyo) was used to measure the ventilatory volume of each breath using a hot-wire flowmeter [38]. Before each exercise testing, we calibrated the test appropriately according to standard protocols.

### Calculation of cardiopulmonary exercise testing measurements

We calculated the difference between inspiratory and expiratory tidal volumes (TV I and E, respectively) for each breathing from breath-by-breath data in each CPET [37]. Further, we have defined the difference between the expiratory and inspiratory tidal volumes to “TV E-I.” We plotted TV E-I against the time axis. In addition, we calculated the mean and standard deviation of TV E-I per minute based on the start of the W.U (zero).

We extracted other CPET parameters, such as the VE/CO_2_ slope and the Y-intercept, minimum VE/VCO_2_, VO_2_/HR, and dead-space gas volume to tidal volume ratio (VD/VT) for each case.

### Spirometry testing

Spirometry testing was performed following standard methods using respiratory function testing equipment (mainly electronic diagnostic spirometer Spiroshift SP-770 COPD Fukuda Denshi Co., Tokyo).

### Statistical analysis

Data were presented as mean ± standard deviation (SD) and 95% confidence intervals. In addition, unpaired data were analyzed using the Student’s t-test. In contrast, paired data were analyzed using a paired t-test. Furthermore, plots of FEV1.0% and Y-intercept of VE/VCO_2_ slope, the difference between minimum VE/VCO_2_ and VE/VCO_2_ slope were linearly regressed, and regression equations and coefficients were calculated. Statistical analyses were performed using Statistics for Excel 2012 (Social Survey Research Information Co., Tokyo, Japan).

### Ethical considerations

The study was conducted following the principles outlined in the Declaration of Helsinki and approved by the ethical committees of Hakodate National Hospital (approval number: R4-0314001) and Gunma Prefectural Cardiovascular Center (approval number:2022020).

The data obtained were delinked and anonymized, and this study was conducted using the data for analysis, with due consideration for protecting the participants’ personal information. The authors confirmed that none of the participants could be identified, and they were fully anonymized. Furthermore, the authors affirmed that all mandatory health and safety procedures complied with the course of conducting the experimental work reported in this paper.

## Results

There were no differences in physical parameters between the convex and non-convex groups; however, there were significantly more males in the convex group. Although there was no significant difference in smoking history between the two groups (Smoking history (+:-) Total 339:195, convex 85:44, non-convex 254:151, p= 0.514), the convex group had more severe cases when the GOLD classification was applied (GOLD classification (0:I:II:III:IV); total 397:86:39:9:3, convex 79:24:15:8:3, non-convex 318:62:24:1:0, p<0.001) (Table 1).

**Table 1.**
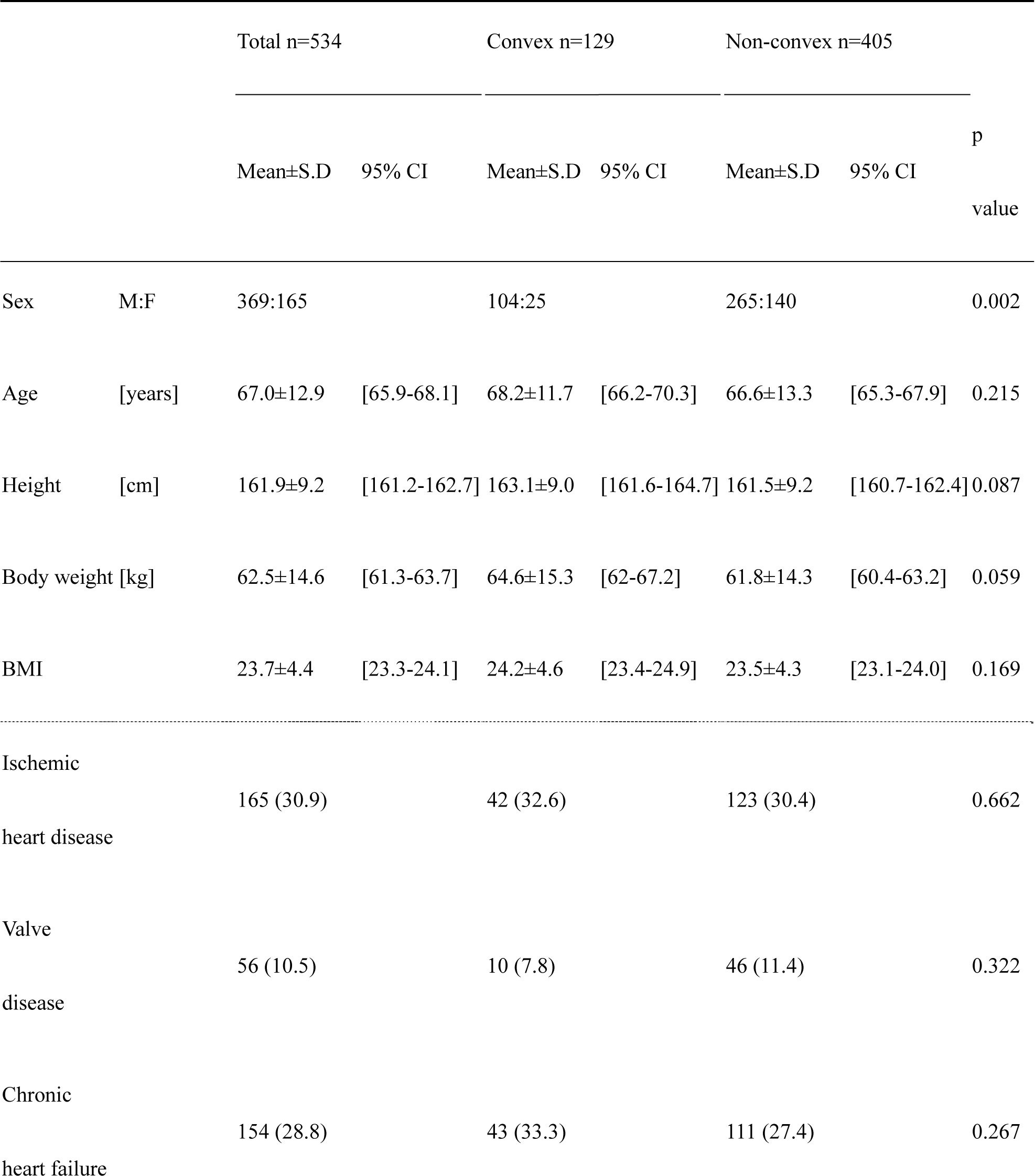
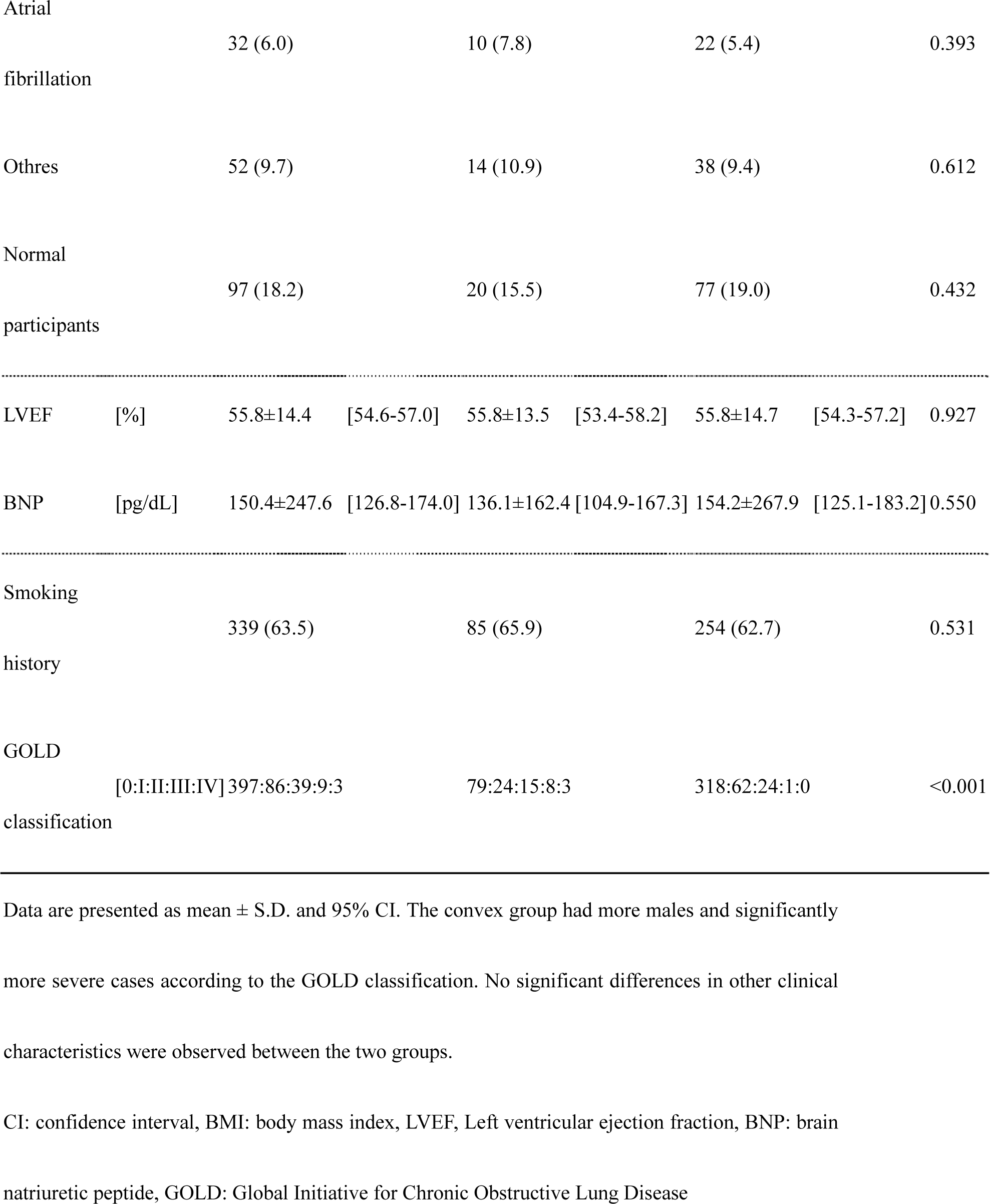
Data of participants’ clinical characteristics

Furthermore, all the participants underwent CPET for symptomatic maximal. Moreover, there were 129 patients in the convex group with decreased TV E-I during CPET and 405 patients in the non-convex group. Although the rating of perceived exertion at the end of the exercise test showed no difference in shortness of breath between the two groups, lower extremity fatigue was significantly lower in the convex group.

### The indices of cardiopulmonary exercise testing

A list of typical CPET parameters is shown in Table 2. There were no significant differences in the exercise tolerance indices between the two groups. Although there were no differences in the VE/VCO_2_ slope and Y-intercept between the two groups, the minimum VE/VCO_2_ tended to be higher in the convex group.

**Table 2.**
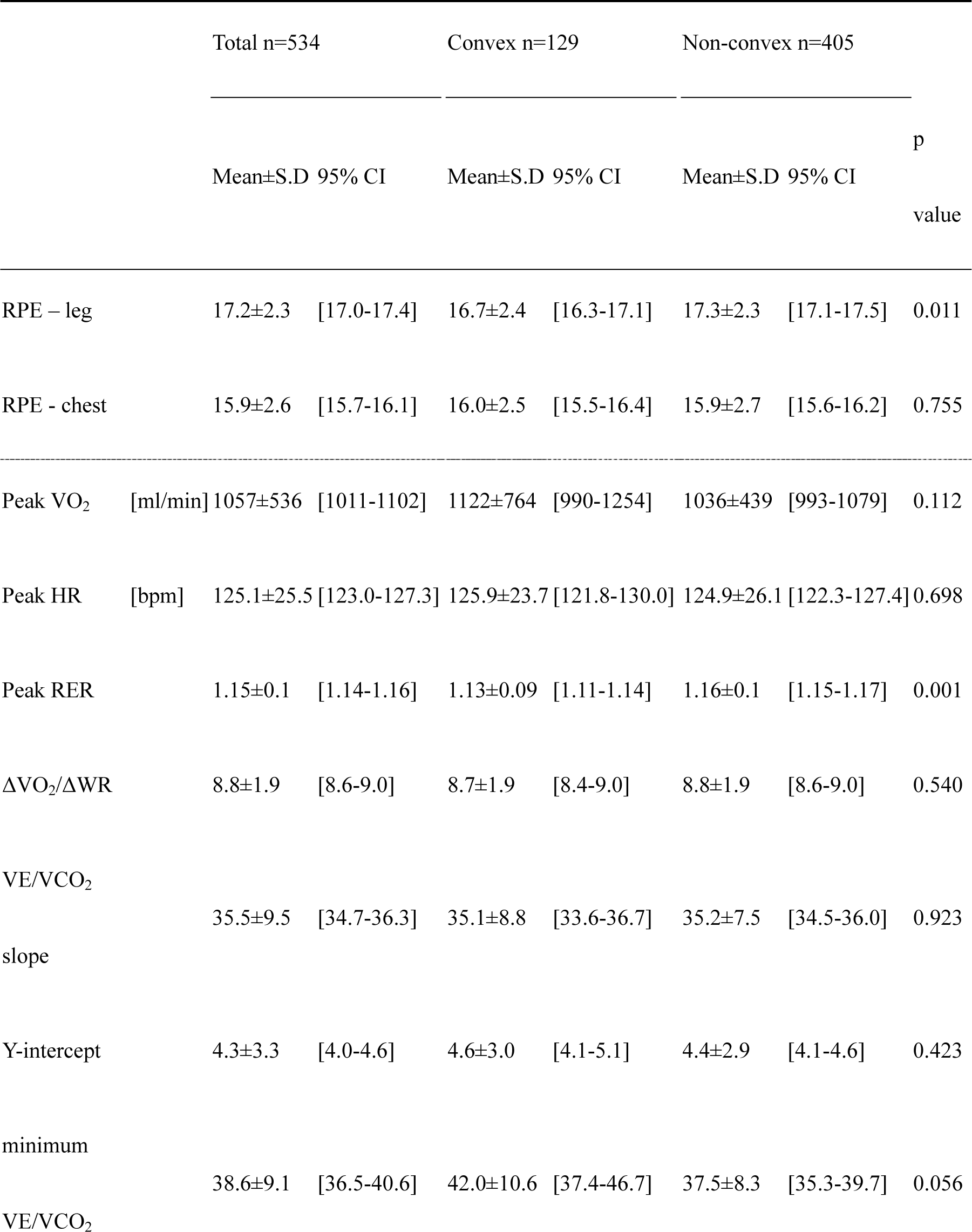
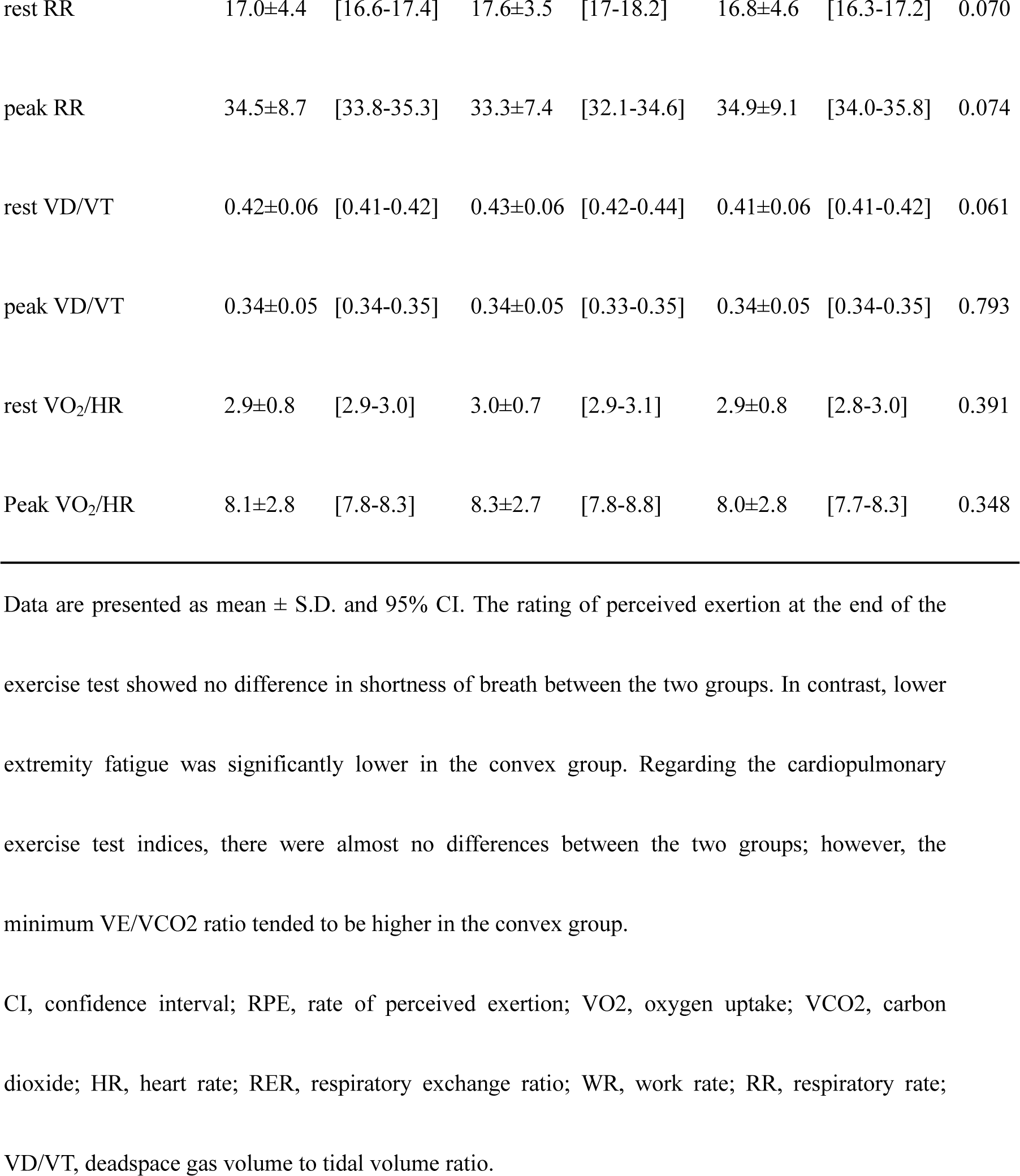
Cardiopulmonary exercise testing indices

### The indices of spirometry testing

Ventilatory capacity (VC), tidal volume (TV), expiratory reverse volume (ERV) and inspiratory reverse volume (IRV) did not differ significantly. However, the convex group had significantly lower FEV1.0% and predictive rates for %FEV1.0, %PEF, and %MVV.

### Relationship between exhaled gas analysis index and spirometry testing

In the convex group, Y-intercept and FEV1.0% showed a significant negative correlation (convex; r=−0.343 [−0.487 ≦ρ≦ −0.181], p<0.001, non-convex; r=−0.090 [−0.186 ≦ρ≦ 0.008], p=0.070) (Figure 1 (A-1, 2)).

**Figure 1.**
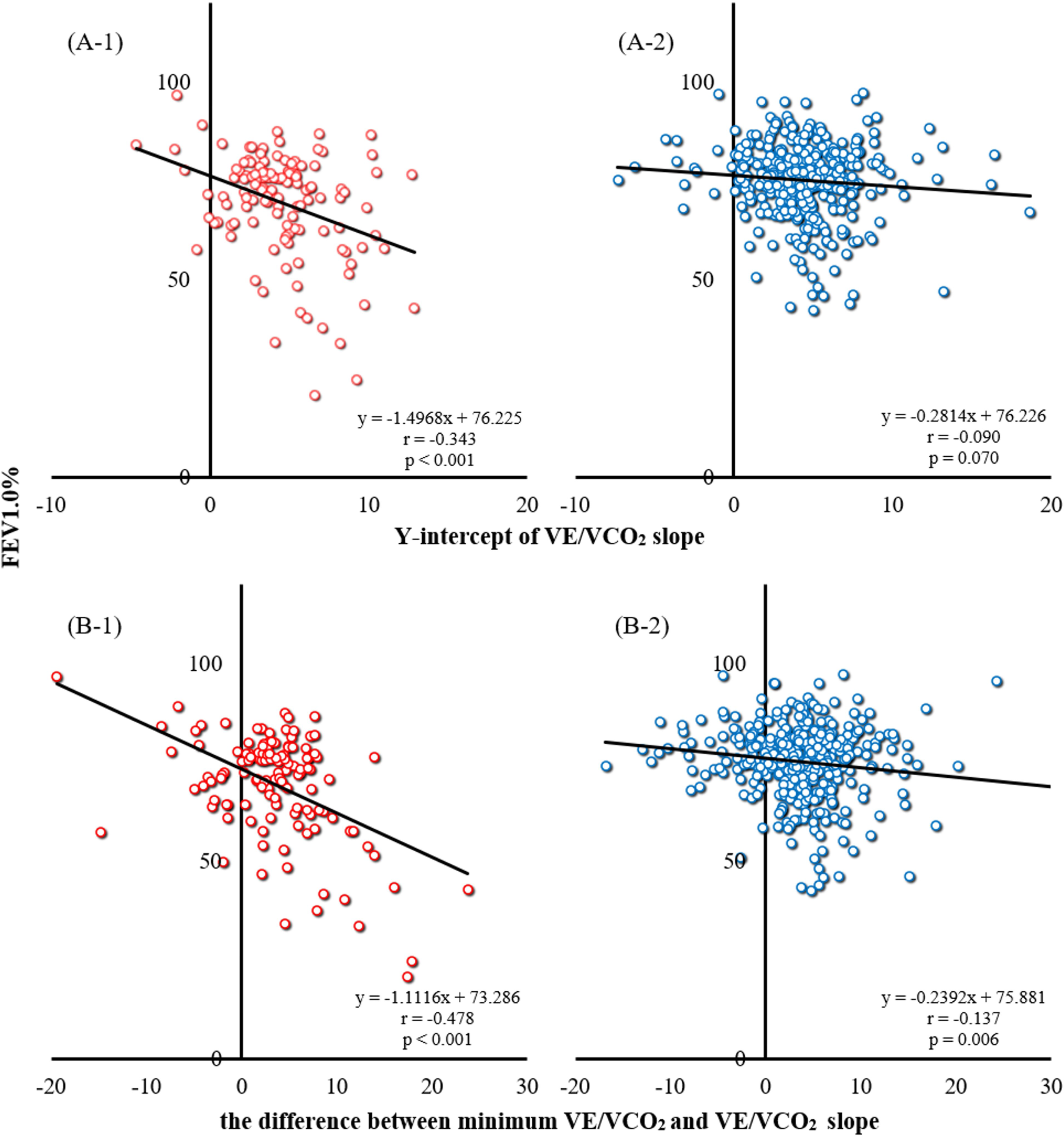
Correlation between respiratory function and CPET indies The upper two panels show the relationship between FEV1.0% and the Y-intercept of VE/VCO_2_ in the convex group (A-1) and the non-convex group (A-2). The lower two panels show the relationship between FEV1.0% and the difference between the minimum VE/VCO_2_ and VE/VCO_2_ slopes in the convex group (B-1) and non-convex groups (B-2).

VE/VCO_2_ slope and minimum VE/CCO_2_ showed little correlation with FEV1.0%; however, there was a significant negative correlation with the difference between VE/VCO_2_ slope and minimum VE/VCO_2_ (convex; r=−0.478 [−0.601≦ρ≦−0.333], p<0.001, non-convex; r=−0.137 [−0.231≦ρ≦−0.040], p=0.006) (Figure 1 (B-1, 2)).

**Table 3.** Spirometry testing indices

|  |  | Total n=534 |  | Convex n=129 |  | Non-convex n=405 |  | p value |
| --- | --- | --- | --- | --- | --- | --- | --- | --- |
|  |  | Mean±S.D | 95% CI | Mean±S.D | 95% CI | Mean±S.D | 95% CI |  |
| VC | [L] | 3.12±0.84 | [3.05-3.19] | 3.18±0.81 | [3.04-3.32] | 3.10±0.86 | [3.01-3.18] | 0.363 |
| %VC | [%] | 103.2±18.7 | [101.6-104.8] | 104.6±18.6 | [101.3-107.8] | 105.6±18.7 | [103.8-107.4] | 0.573 |
| TV | [L] | 0.91±0.37 | [0.87-0.94] | 0.94±0.45 | [0.86-1.02] | 0.90±0.34 | [0.86-0.93] | 0.246 |
| ERV | [L] | 0.96±0.48 | [0.92-1.00] | 0.95±0.44 | [0.88-1.03] | 0.96±0.49 | [0.91-1.01] | 0.872 |
| %ERV | [%] | 76.5±34.0 | [73.6-79.3] | 72.6±31.3 | [67.2-78.0] | 77.7±34.8 | [74.3-81.1] | 0.142 |
| IRV | [L] | 1.25±0.58 | [1.20-1.30] | 1.28±0.54 | [1.19-1.37] | 1.24±0.59 | [1.18-1.30] | 0.511 |
| IC | [L] | 2.16±0.64 | [2.10-2.21] | 2.22±0.63 | [2.11-2.33] | 2.14±0.64 | [2.07-2.20] | 0.207 |
| FVC | [L] | 3.06±0.85 | [2.98-3.13] | 3.11±0.83 | [2.97-3.25] | 3.04±0.85 | [2.95-3.12] | 0.395 |
| %FVC | [%] | 92.4±16.6 | [91.0-93.8] | 91.4±17.3 | [88.5-94.4] | 92.7±16.3 | [91.1-94.3] | 0.451 |
| FEV1.0 | [L] | 2.26±0.72 | [2.20-2.32] | 2.17±0.74 | [2.05-2.30] | 2.28±0.71 | [2.21-2.35] | 0.130 |
| %FEV1.0 | [%] | 101.5±23.4 | [99.5-103.4] | 96.2±28.0 | [91.4-101.0] | 103.1±21.5 | [101.0-105.2] | 0.003 |
| FEV1.0% | [%] | 73.7±10.4 | [72.8-74.6] | 69.4±13.1 | [67.1-71.6] | 75.0±9.0 | [74.2-75.9] | 0.000 |
| MMF | [L/s] | 2.02±1.06 | [1.78-2.27] | 1.64±0.83 | [1.26-2.03] | 2.10±1.06 | [1.82-2.38] | 0.100 |
| %MMF | [%] | 68.7±28.5 | [62.2-75.2] | 58.3±28.1 | [45.3-71.3] | 70.3±25.1 | [63.7-77.0] | 0.091 |
| PEF | [L/s] | 6.33±2.34 | [5.80-6.86] | 5.54±2.15 | [4.60-6.48] | 6.61±2.36 | [5.99-7.24] | 0.079 |
| %PEF | [%] | 83.6±23.1 | [78.4-88.8] | 72.8±24.8 | [61.9-83.7] | 87.5±21.4 | [81.9-93.2] | 0.014 |
| MVV | [L/min] | 68.6±25.7 | [66.1-71.0] | 74.8±30.3 | [69-80.6] | 81.0±28.3 | [77.9-84.0] | 0.058 |
| %MVV | [%] | 84.7±24.7 | [82.4-87.0] | 92.2±31.2 | [86.2-98.2] | 103.2±27.1 | [100.3-106.1] | 0.001 |

Data are presented as mean ± S.D. and 95% CI. The measured values of forced expiratory volume in 1 s as a percentage of forced vital capacity (FEV1.0%), predicted values of forced expiratory volume in 1 s (%FEV1.0), peak expiratory flow (%PEF), and maximal ventilatory volume (%MVV) were significantly lower in the convex group.
CI, confidence interval; VC, vital capacity; VT, tidal volume; ERV, expiratory reserve volume; IRV, inspiratory reserve volume; IC, inspiratory capacity; FVC, forced vital capacity; FEV, forced expiratory volume; FEV1.0, forced expiratory volume in 1 s; FEV1.0%, forced expiratory volume in 1 s as a percentage of forced vital capacity; MMF, maximal mid-expiratory flow; PEF, peak expiratory flow; MVV, maximal ventilatory volume.

## Discussion

This is the first study to examine the relationship between differences in respiratory function and CPET indices based on the qualitative detection of DLH using CPET. DLH is a physiological respiratory mechanism in which expiration occurs less than inspiration, with an increased respiratory rate. Based on this theory, we showed in this study that DLH could be easily detected and correlated with respiratory function by analyzing data from exhaled gas analyses.

### Respiratory function by spirometry testing

Respiratory functions, such as VC and TV, did not differ between the two groups; however, FEV1.0%, %MVV, and %PEF were lower in the convex group. In the convex group, the mean value of FEV1.0% was equivalent to the diagnostic criteria for obstructive ventilatory impairment. In contrast, in the non-convex group, most cases did not meet the criteria for GOLD stage 1, and several cases were not diagnosed as having obstructive ventilation impairment on spirometry testing. Nevertheless, several patients in the convex group likely had DLH because of peripheral airway obstruction (stenosis) or other factors, as they had less expiration than inspiration during the exercise. Therefore, it is possible that respiratory function had already declined before the criteria for diagnosis of DLH because of the obstruction of the small bronchioles and other organs. Furthermore, it has already been shown that subclinical respiratory impairment may also affect cardiovascular function and cardiovascular disease. Although this was a cross-sectional study and the outcome of very mild cases is unknown, we believe that TV E-I may be a qualitative indicator of peripheral airway obstruction.

### Relationship between cardiopulmonary exercise testing indices and dynamic pulmonary hyperinflation

Ventilatory efficiency decreases because of congestion caused by heart failure and obstructive ventilation impairment [39]. For instance, in patients with heart failure, the minimum VE/VCO_2_ and VE/VCO_2_ slope increase with disease severity; however, they are generally consistent [40]. In contrast, in COPD, the VE/VCO_2_ slope increases in mild disease but decreases in severe disease [41, 42]. Furthermore, it has been reported that in COPD, the Y-intercept of the VE/VCO_2_ slope is related to FEV1.0%, and the Y-intercept is higher [36]. Murata et al. reported that as COPD progresses, the minimum VE/VCO_2_ and VE/VCO_2_ slopes may diverge, or the VE/VCO_2_ slope may become pseudo-negative, and the Y-intercept may be high [36, 43]. However, both indices only report observational studies on COPD and do not examine the presence or absence of DLH or its extent.

Although the convex group showed a trend toward a higher minimum VE/VCO_2_ in this study, there were no significant differences in these indices between the two groups, including the Y-intercept and VE/VCO_2_ slope. However, even in such cases, FEV1.0% and the Y-intercept of the VE/VCO_2_ slope showed a significant correlation in the convex group, similar to the results of previous studies. Furthermore, FEV1.0% also showed a significant negative correlation with the difference between the minimum VE/VCO_2_ ratio and the VE/VCO_2_ slope. In contrast, in the non-convex group, the correlation between the difference in minimum VE/VCO_2_ and VE/VCO_2_ slope and FEV1.0% was very limited, indicating that a combined evaluation with TV E-I is crucial.

Although the study group had milder respiratory function impairment than those in previous studies, it was suggested that the combined TV E-I and Y-intercept or the difference between the minimum VE/VCO_2_ and VE/VCO_2_ slope during incremental exercise testing could provide an index of respiratory function and an assessment of the severity of peripheral airway obstruction in patients with stable cardiac disease.

### The usefulness of detecting dynamic lung hyperinflation using cardiopulmonary exercise test

Most participants in this study had stable heart disease and mild respiratory function impairment. However, 63.4% of the patients had a smoking history, and more than 20% showed the possibility of DLH on CPET. Interestingly, the prevalence of COPD is expected to decrease in high-income countries as smoking declines; however, it will become a major social problem in low-to-middle-income countries [44]. Thus, COPD causes chronic systemic inflammation, leading to a decline in physical function and a worsening prognosis [45]. Moreover, DLH increases the respiratory workload and restricts venous return [46], leading to exercise limitation and static lung hyperinflation caused by COPD progression and a worsened prognosis [45,47,48].

We believe that capturing respiratory changes associated with increased exercise intensity using CPET, as in this study, is a simple and qualitative method for detecting airway stenosis and DLH at an early stage, even in patients with mild or asymptomatic symptoms. Furthermore, we believe this will lead to early scrutiny, appropriate therapeutic interventions, and drug prescriptions, thus, improving the quality of life and patient prognosis.

### Limitations

There are several limitations to this study.

First, this was a cross-sectional study, and cardiopulmonary exercise and spirometry testing were not often performed at approximately the same time, possibly resulting in a selection bias (comorbidity of DLH). In addition, it is unclear about the course of the subjects in this study, whether there was worsening shortness of breath and other factors, progressive decline in respiratory function, or a diagnosis of COPD. Therefore, further studies are warranted.

Second, the study did not compare the results with those of the existing DLH assessment methods or evaluate the response to bronchodilator use. Therefore, it is difficult to confirm the presence of DLH based on the results of this study alone.

Third, several participants in this study had relatively preserved respiratory function. Therefore, it is uncertain whether a similar trend would be observed in patients with moderate-to-severe COPD who have already been diagnosed.

Finally, it is currently difficult to determine the severity of DLH; therefore, developing appropriate analytical methods for TV E-I is desirable.

## Conclusion

Evaluating data on differences in expiratory and inspiratory tidal volumes (TV E-I) during cardiopulmonary exercise testing has proven useful for dynamic lung hyperinflation in patients with stable heart disease. The combined evaluation of the TV E-I and Y-intercept of the VE/VCO_2_ slope, or the difference between the minimum VE/VCO_2_ and VE/VCO_2_ slopes in CPET, could detect cases of potential respiratory impairment or peripheral airway obstruction.

## Data Availability

The dataset used in this study is available from the corresponding author upon request.

## Abbreviations

VAT: ventilatory anaerobic threshold
Inc-Ex: incremental exercise
CPET: Cardiopulmonary exercise testing
VO2: Oxygen uptake
VCO2: carbon dioxide production
VE: ventilatory equivalent
RPE: Rating of perceived exertion
RR: respiratory rate
VD/VT: dead-space gas volume to tidal volume ratio
COPD: chronic obstructive pulmonary disease
DLH: dynamic lung hyperinflation
TV: tidal volume
TV I: inspiratory tidal volume
TV E: expiratory tidal volume
FEV: forced expiratory volume
FEV1.0: forced expiratory volume in 1 second
FEV 1.0%: forced expiratory volume in 1 second as a percent of forced vital capacity
VC: vital capacity
IC: inspiratory capacity

## Acknowledgements

We would like to thank Editage for assistance with English language editing.

The results of the study are presented clearly, honestly, and without fabrication, falsification, or inappropriate data manipulation, and the results of the present study do not constitute an endorsement by the *Journal of American Heart Association*.

## Authors’ contributions

KK, MM, and MA developed the study concept and were involved in its design and implementation. KK, KN, NM, and MM delivered program content to the participants. KK, KN, NM, MU, YK, and MM acquired data. KK analyzed the data. KK and MM prepared the manuscript. KN, MN, KY, and MA drafted the manuscript and approved the final draft. All the authors have read and approved the final version of the manuscript.

## Availability of data and materials

The dataset used in this study is available from the corresponding author upon request.

## Ethics approval and consent to participate

The study was conducted following the principles outlined in the Declaration of Helsinki and was approved by the ethics committees of Hakodate National Hospital (approval number: R4-1114006) and the Gunma Prefectural Cardiovascular Center (approval number:2022020).

The data obtained were delinked and anonymized, and this study was conducted using the data for analysis and due consideration to the protection of the participants’ personal information. Consequently, the authors confirmed that none of the participants could be identified, and they were fully anonymized. Furthermore, the authors affirm that all mandatory health and safety procedures complied with the course of conducting the experimental work reported in this paper.

## Consent for publication

Not applicable.

